# Clustered Mendelian Randomization analyses identifies distinct and opposing pathways in the causal association between insulin-like growth factor-1 and type 2 diabetes mellitus

**DOI:** 10.1101/2021.05.12.21257093

**Authors:** Wenyi Wang, Ephrem Baraki Tesfay, Ko Willems van Dijk, Andrzej Bartke, Diana van Heemst, Raymond Noordam

## Abstract

**Aims/hypothesis:** There is inconsistent evidence for the causal role of serum insulin-like growth factor-1 (IGF-1) concentration in the pathogenesis of type 2 diabetes. Here, we investigated the association between IGF-1 and type 2 diabetes using a combination of multivariable-adjusted and (clustered) Mendelian Randomization (MR) analyses in the UK Biobank.

**Methods:** We conducted Cox proportional hazard analyses in 451,232 European-ancestry individuals of the UK Biobank (55.3% women, mean age at recruitment 56.6 years), among which 13,247 individuals developed type 2 diabetes during up to 12 years of follow-up. In addition, we conducted two-sample MR analyses based on independent SNPs associated with IGF-1. Given the heterogeneity between the causal estimates of individual instruments (P-value for Q statistic=4.03e-145), we also conducted clustered MR analyses. Biological pathway analyses of the identified clusters were performed by overrepresentation analyses.

**Results:** In the Cox proportional hazard models, with IGF-1 concentrations stratified in quintiles, we observed that participants in the lowest quintile had the highest relative risk of type 2 diabetes (HR: 1.31; CI: 1.23-1.39). In contrast, in the two-sample MR analyses, higher genetically-influenced IGF-1 was associated with a higher risk of type 2 diabetes. Based on the heterogeneous distribution of causal effect estimates, six clusters associated either with a lower or a higher risk of type 2 diabetes were identified. The main clusters in which a higher IGF-1 was associated with a lower risk of type 2 diabetes consisted of instruments mapping to genes in the growth-hormone signaling pathway, whereas the main clusters in which a higher IGF-1 was associated with a higher risk of type 2 diabetes consisted of instruments mapping to genes in pathways related to amino acid metabolism and genomic integrity.

**Conclusion:** The IGF-1 associated SNPs used as genetic instruments in MR analyses showed a heterogeneous distribution of causal effect estimates on the risk of type 2 diabetes. This was likely explained by differences in the underlying molecular pathways that increase IGF-1 concentration and differentially mediate the effects of IGF-1 on type 2 diabetes.

## Introduction

Insulin-like growth factor-1 (IGF-1) is a pleiotropic hormone that plays a major role in cellular growth, proliferation and survival [1]. The secretion of IGF-1, predominately by the liver, is promoted by growth hormone (GH) while conversely, IGF-1 in the circulation feeds back centrally to hypothalamus to inhibit GH secretion [2]. The availability of free IGF-1 in the blood is regulated by its association with distinct insulin-like growth factor binding proteins (IGFBPs) which can increase IGF-1 half-life or block its binding to IGF-1 receptors [3]. IGF-1 was found to be involved in the pathophysiology of various diseases, including cancer, neurodegenerative disease, cardiovascular disease, and type 2 diabetes mellitus [4]. Several (prospective) cohort studies have found that lower levels of IGF-1 were associated with an increased risk of impaired glucose tolerance, increased insulin resistance and hence the development of diabetes mellitus [5, 6].

In contrast to these multivariable-adjusted association analyses, a recent study demonstrated that a higher genetically-influenced IGF-1 concentration was associated with a higher risk of developing type 2 diabetes using Mendelian Randomization ([0-9]+)(MR) analyses [7]. MR is an approach to determine whether the association between risk factors and outcome is causal by using genetic variants as instrumental variables [8]. However, in some circumstances, there is clear heterogeneity in the causal effects of the individual single nucleotide polymorphisms (SNPs) that are used as instrumental variables, which may indicate either pleiotropy or differences in biological pathways contributing to high levels of the exposure [9]. Clustered MR was recently developed to provide a means to address the heterogeneity in causal effects by clustering variants that show similar individual causal estimates on the outcome [9]. Previously, such context-dependent MR analyses have been proposed to provide more biological perspective in causal associations [10, 11].

We hypothesized that heterogeneity in causal effects of individual variants could be a reflection of different biological mechanisms involved in the association between IGF-1 and type 2 diabetes. For example, variation influenced by processes causing insufficient GH signaling may have a different impact on T2D than variation influenced by processes causing increased GH resistance. Therefore, in this study, we aimed to investigate the association between IGF-1 and incident type 2 diabetes followed by clustered MR analyses [9] in the large UK Biobank population, and explored the possible biological pathways involved in the clustered causal associations.

## Methods

### Study setting and study population

The UK Biobank is a very large prospective cohort study with over 500,000 participants aged 40-69 years at recruitment across the entire United Kingdom (UK) [12]. Participants were recruited between 2006 and 2010 in 22 assessment centers across the UK. Baseline examinations in all participants included physical measures, collection of blood, urine and saliva, a self-completed touch-screen questionnaire, and a brief computer-assisted interview to investigate sociodemographic, family history, environmental factors, lifestyle, psychosocial factors, etc. The UK Biobank study was approved by the North-West Multi-center Research Ethics Committee (MREC). Access to information to invite participants was approved by the Patient Information Advisory Group (PIAG) from England and Wales. All participants in the UK Biobank study provided written informed consent.

The present project was accepted under project number 22474. We restricted the analyses to the UK Biobank participants who reported to be of European ancestry including British, Irish and any other European background, who had information available on serum IGF-1 concentration, and who were in the full release imputed genomics datasets.

### Genotyping and genetic imputations

Genome-wide genotype data for all 500,000 UK Biobank participants generated using Affymetrix UK BiLEVE Axiom array (initial 50,000 participants) and the Affymetrix UK Biobank Axiom Array (remaining 450,000 participants), which genotyped around 850,000 variants. All genetic data were quality controlled centrally by UK Biobank resources. In addition, UK Biobank resources performed centralized imputations on approximately 96 million genotypes using the UK10K haplotype [13], 1000 Genomes Phase 3 [14], and Haplotype Reference Consortium (HRC) reference panels [15]. Autosomal SNPs were pre-phased using SHAPEIT3 and imputed using IMPUTE4. Related individuals were identified by estimating kinship coefficients for all pairs of samples using only markers weakly informative of ancestral background. More information on the genotyping processes and genetic imputation can be found online (https://biobank.ctsu.ox.ac.uk/crystal/label.cgi?id=263).

### Biochemical analyses

Biological samples were collected to measure biochemical markers including IGF-1 at baseline (2006-2010) comprising ∼480,000 participant samples. Serum levels of IGF-1 were analyzed using chemiluminescent Immunoassay (DiaSorin Liaison XL) with a one-step sandwich. Coefficients of variation derived from the internal quality control samples of the low, medium, and high IGF-1 concentrations ranged from 6.03-6.18%. More information on assay performance of the UK Biobank Biomarker Project can be found online (https://biobank.ctsu.ox.ac.uk/crystal/refer.cgi?id=1227).

### Outcome definitions

Information on the diagnosis of type 2 diabetes during follow-up was obtained through linkage with the National Health System (NHS) medical records database. Diagnoses were mainly derived from hospital admissions data. More information related to the diagnosis of type 2 diabetes can be found on the websites of UK Biobank under data-field 130708 (https://biobank.ctsu.ox.ac.uk/crystal/field.cgi?id=130708).

### Prospective analyses

Participants without diabetes mellitus at baseline were followed until the occurrence of type 2 diabetes, mortality or loss of follow-up, whichever occurred first. The association between IGF-1 levels and incidence of type 2 diabetes in the UK Biobank cohort was assessed using Cox proportional hazard models. Participants were categorized into five groups based on quintiles of IGF-1 concentration. Quintile 1 (lowest 20%) and quintile 5 (highest 20%) were used separately as reference groups to calculate the hazard ratio (HR), respectively. Potential confounders included sex, age at recruitment and baseline body mass index (BMI) based on height and weight measured at the assessment centers. The analyses were conducted in R using the survival package (version 3.2-7) [16]. The Kaplan Meier curve was plotted to visualize the difference of survival probability between IGF-1 quintiles and whether the proportionality assumption holds.

### Genome-wide association analyses

Genome-wide association analyses (GWAS) on continuous IGF-1 concentrations and dichotomized IGF-1 blood levels (lowest 20% versus remaining 80% and highest 20% versus remaining 80%) were performed to provide a list of independent lead SNPs to be used as instrumental variables in the MR analyses. Analyses were performed using linear mixed models implemented in the program BOLT_LMM (version 2.3.2) [17]. We adjusted the analyses for age, sex and the first 10 principal components, and corrected for the Kinship matrix to correct for familial relationships in the UK Biobank population. Analyses were done on the autosomal chromosomes only. SNPs with a minor allele frequency <0.01 as well as SNPs with an imputation quality <0.3 were excluded. P-values of SNPs smaller than 5e^-8^ were extracted and stored for the MR analyses. Visualization of the results was performed using the R-based packages ggplot2 [18] and EasyStrata [19] (www.genepi-regensburg.de/easystrata).

### Mendelian randomization analyses

Two-sample MR was performed with summary-based statistics of GWAS using the R-based statistical package Two Sample MR (available at: http://github.com/MRCIEU/TwoSampleMR) [20]. This statistical package also connects to a large library of exposures from published GWAS to use as instrumental variables, which is aligned with the online GWAS catalogue.

For the present study, we performed the clumping process (window size = 10,000 kb, R^2^ < 0.001) with the European samples from the 1000 genomes project which were used to estimate linkage disequilibrium (LD) between SNPs. Among those pairs of SNPs with R^2^ above the specified threshold (R^2^ = 0.001), only the SNPs with the lowest P-value were retained to provide a list of independent lead SNPs from the MR analyses. Otherwise, the statistical power of MR analyses would be overestimated (e.g., underestimated standard errors of the summary estimates of the MR analyses). SNPs present in UK Biobank, but absent from the LD reference panel, were removed. On the basis of the significant independent lead SNPs (P-value<5e-8), we assessed their possible causal association with type 2 diabetes. For this analysis, we used publicly-available summary-statistics GWAS meta-analysis from 32 studies on 898,130 individuals (74,124 type 2 diabetes cases and 824,006 controls) of European ancestry [21].

Methods for MR analyses of summary-level data based on a two-sample design have been described in detail previously [22, 23]. Using inverse-variance-weighted (IVW) analyses, we combined the effects of the individual genetic instruments to obtain a genetically-determined association between exposure and outcome under the assumption of the absence of horizontal pleiotropy. However, given the large number of genetic instruments included in the present analyses, there is a high probability that at least some SNPs show pleiotropic effect. To test whether possible pleiotropic effects could bias the overall effect estimates (horizontal pleiotropy), we performed the sensitivity analyses MR Egger regression [24] and weighted median estimator (WME) analyses [25]. MR Egger does not force the regression line through the intercept and is, therefore, able to test for the presence of directional pleiotropy, and WME estimator assumes at least 50% of the instruments included in the MR analyses were valid.

### Clustered Mendelian randomization analyses

Clustered MR analyses were conducted to identify groups of genetic variants that have similar causal estimates of the exposure on the outcome. If the causal estimates of each genetic variant on the outcome were similar (e.g., their ratio-estimates were similar in direction, magnitude and precision), it was divided into different clusters in which the included genetic variants were more homogeneous concerning the causal estimates [9]. The inclusion probability of SNPs in each cluster was higher than 0.8.

In order to identify distinct causal effects of genetic variants derived from GWAS of continuous IGF-1 levels on type 2 diabetes, we made use of the R-based MR-Clust package (https://github.com/cnfoley/mrclust). MR-Clust performs likelihood-based clustering on Wald ratio-estimates and accompanied standard errors. Genetic instruments within a cluster share similar causal estimates (e.g., Wald ratio-estimates are similar in direction, magnitude and precision) of the causal effect of the exposure on the outcome. MR analyses were repeated on all clusters to investigate the causal effects of each cluster on type 2 diabetes.

### Pathway analyses

Based on the instruments that were part of a certain cluster, we performed pathway analyses to provide insights into biological pathways which could explain the heterogeneity between causal estimates of genetic variants in different clusters on type 2 diabetes. Genetic variants in each cluster were extracted and processed using the online tool FUMA to perform gene mapping where independent lead SNPs were determined at R^2^ < 0.1 using the 1000 Genome Project Phase 3 as a reference panel population [14, 26]. Based on derived genes, pathway analyses were conducted by using GENE2FUNC in the online tool FUMA, which is able to check pathways coded by related genetic variants [27]. The GENE2FUNC connects to massive pathway databases by using overrepresentation enrichment tests, among which KEGG and Reactome databases were used in the pathway analyses. More information can be found on the website of FUMA (https://fuma.ctglab.nl/)

## Results

### Characteristics of the study population

In total, 451,232 European participants without diabetes mellitus at baseline were included in our study of whom 13,247 developed type 2 diabetes in up to 12 years of follow-up (**Table 1**). Of the participants not developing type 2 diabetes, 55.7% were women, the mean age at recruitment was 56.5 (SD=8.1) years, the average BMI was 27.1 (SD=4.5) kg/m^2^ and the mean IGF-1 levels were 21.5 (SD=5.6) nmol/L. Of the participants developing type 2 diabetes during follow-up, women accounted for 41.9%, the mean age at recruitment was 59.3 (SD=7.2) years, the average BMI was 31.7 (SD=5.6) kg/m^2^ and the mean IGF-1 level was 19.9 (SD=6.5) nmol/L.

**Table 1.**
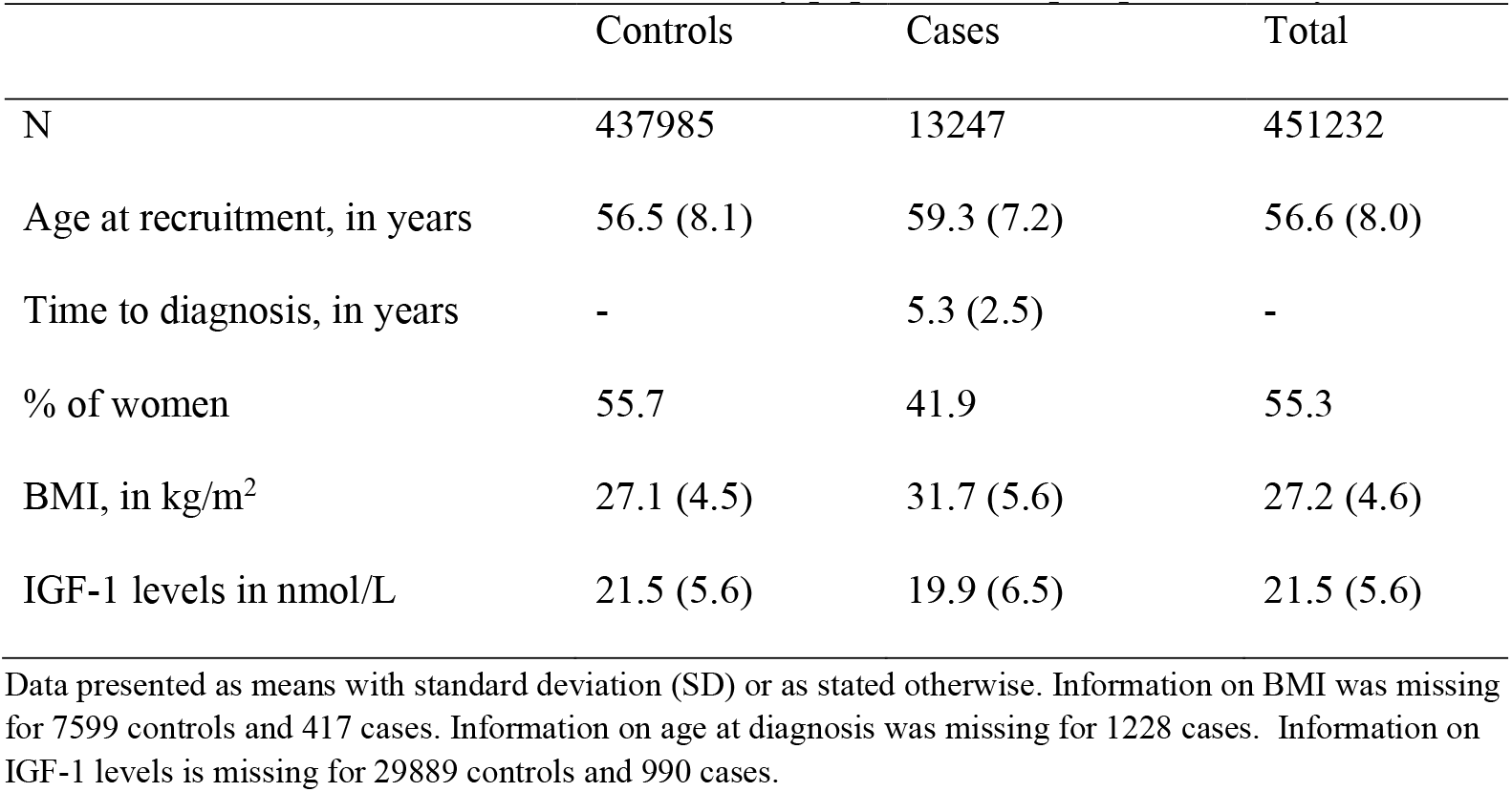
Characteristics of the UK Biobank study population for prospective analyses

### Prospective analyses

Multivariable-adjusted Cox proportional hazard model analyses were performed to evaluate the association between quintiles of IGF-1 levels and incident type 2 diabetes.

Kaplan Meier curve (**Figure 1**) illustrated that participants in the lower IGF-1 quintiles (quintile 1and 2) had proportionally lower risk of type 2 diabetes compared to participants in the higher quintiles (quintile 3, 4 and 5).

**Figure 1.**
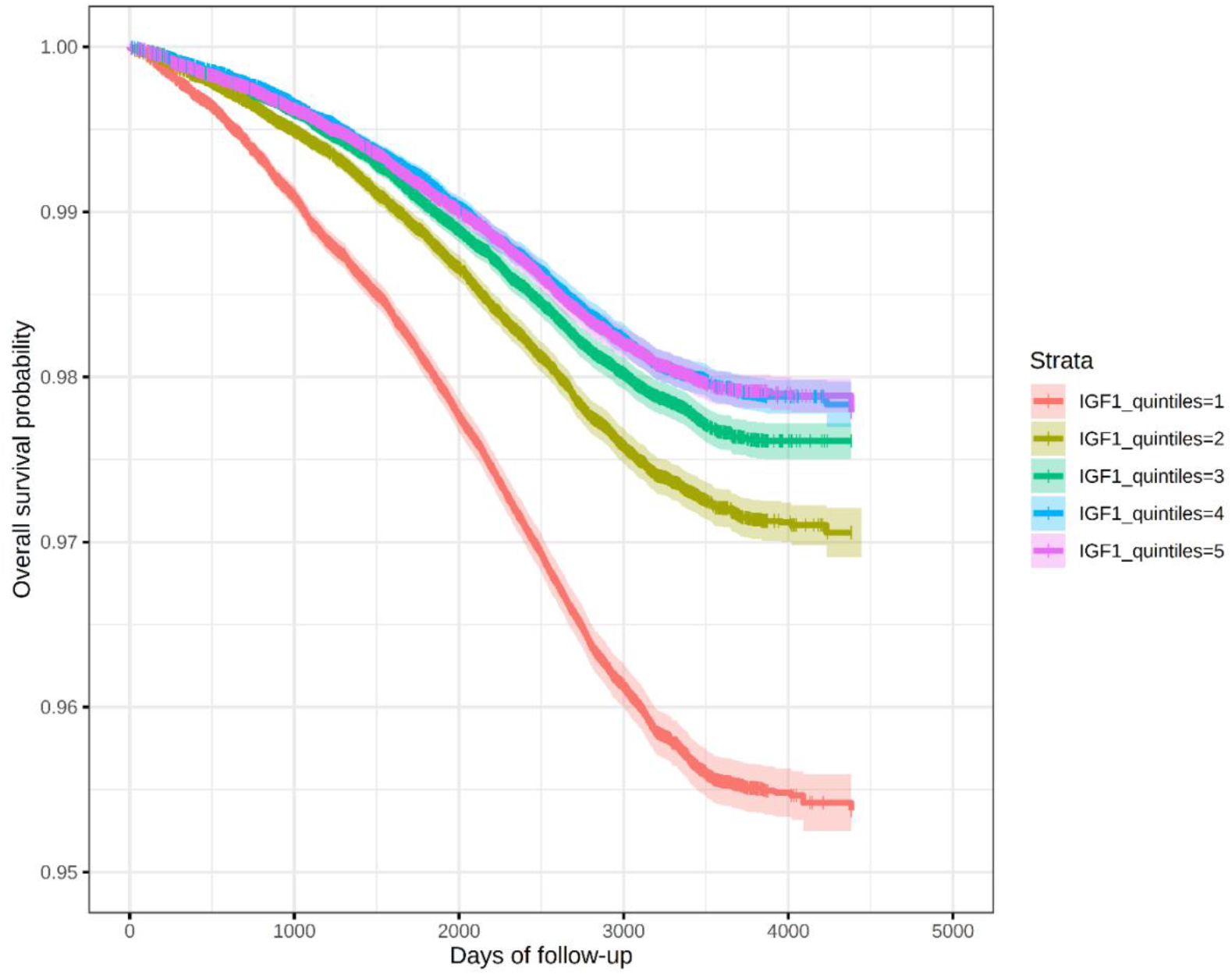
Kaplan Meier survival curves displaying the time to develop incident type 2 diabetes by IGF-1 quintile. The x-axis presents days of follow-up and y-axis presents overall survival probability. Censoring is indicated by vertical marks.

More specifically, individuals in quintile 2 had a lower risk of type 2 diabetes (HR: 0.77; CI: 0.73-0.81) than individuals in quintile 1 (**Table 2**), as did those in quintile 3 (HR: 0.70; CI: 0.66-0.74), quintile 4 (HR: 0.68; CI: 0.64-0.72) and quintile 5 (HR: 0.76; CI: 0.72-0.81). Taking participants in quintile 5 as the reference group, a higher risk was observed in quintile 1 (HR: 1.31; CI: 1.23-1.39), and those in quintile 3 (HR: 0.92; CI: 0.86-0.98) and 4 (HR: 0.89; CI: 0.83-0.95) had a lower risk of developing type 2 diabetes.

**Table 2.**
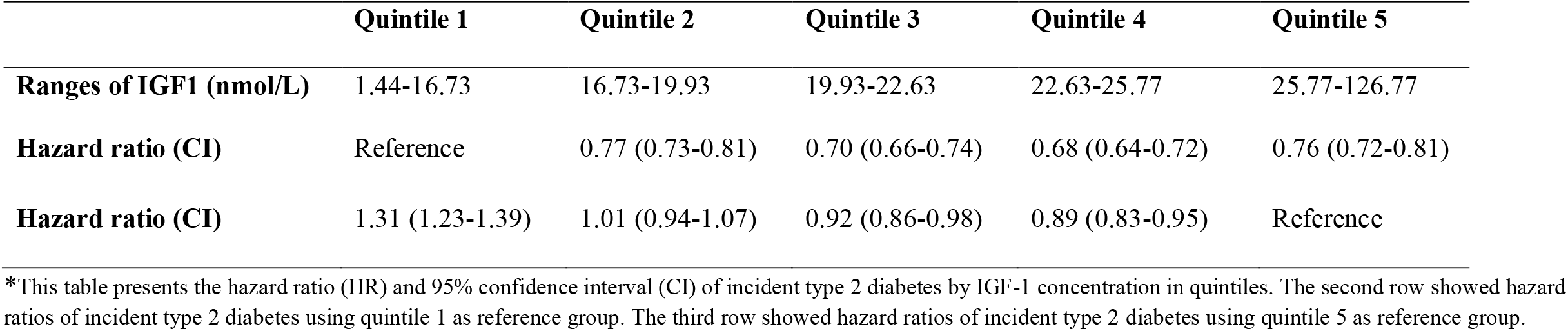
Hazard ratio (HR) with 95% CI of incident type 2 diabetes by IGF-1 concentration in quintiles

### Mendelian Randomization analyses

There were 95,877 significant SNPs (P<5e-8) identified in the GWAS of continuous IGF-1 (**Supplementary Figure 1**) of which we derived 387 independent lead SNPs to be used in the MR analyses. The MR estimates of assessing the causal effect of IGF-1 on type 2 diabetes showed that a 1 nmol/L increase in IGF-1 was associated with a 1% higher risk of type 2 diabetes in the IVW analyses (OR: 1.01; CI: 1.00-1.02). Similar results were obtained from MR Egger (OR: 1.02; CI: 1.00-1.05) and WME analyses (OR: 1.01; CI: 1.00-1.02).

The GWAS on dichotomized IGF-1 for lowest or highest 20% versus the remaining 80% resulted in 154 and 184 independent lead SNPs, respectively. MR analyses showed that low IGF-1 was associated with a lower risk of type 2 diabetes in the IVW analyses (OR: 0.70; CI: 0.53-0.94) and high IGF-1 was associated with a higher risk of type 2 diabetes in the IVW analyses (OR: 1.66; CI: 1.28-2.15) (**Supplementary Table 1, Supplementary Table 2; Supplementary Figure 2)**. Results remained similar in the MR-Egger and WME analyses.

### Clustered Mendelian randomization analyses

We observed large heterogeneity in the individual causal estimates (**Figure 2**) as was also evidenced by the Q statistic (P=4.03e^-145^). The individual causal estimates were clustered into a total of 6 clusters with an inclusion probability of SNPs higher than 0.8 (**Figure 3**). MR estimates from different methods of assessing the causal effect of 6 clusters on type 2 diabetes are presented in **Table 3** and **Supplementary Figure 3**. In cluster 1 (IVW: OR:1.54, CI:1.43-1.65), cluster 2 (IVW: OR:1.03, CI:1.02-1.04) and cluster 5 (IVW: OR:1.20, CI:1.18-1.22), higher levels of IGF-1 level were associated with a higher risk of type 2 diabetes. On the other hand, cluster 3 (IVW: OR:0.92, CI:0.91-0.94), cluster 4 (IVW: OR:0.62, CI:0.58-0.67) and cluster 6 (IVW: OR:0.80, CI:0.76-0.85) showed that higher levels of IGF-1 were associated with lower risk of type 2 diabetes. The results from sensitivity analyses by using weighted median estimator and MR-Egger did not materially differ from the result of the IWV method.

**Figure 2.**
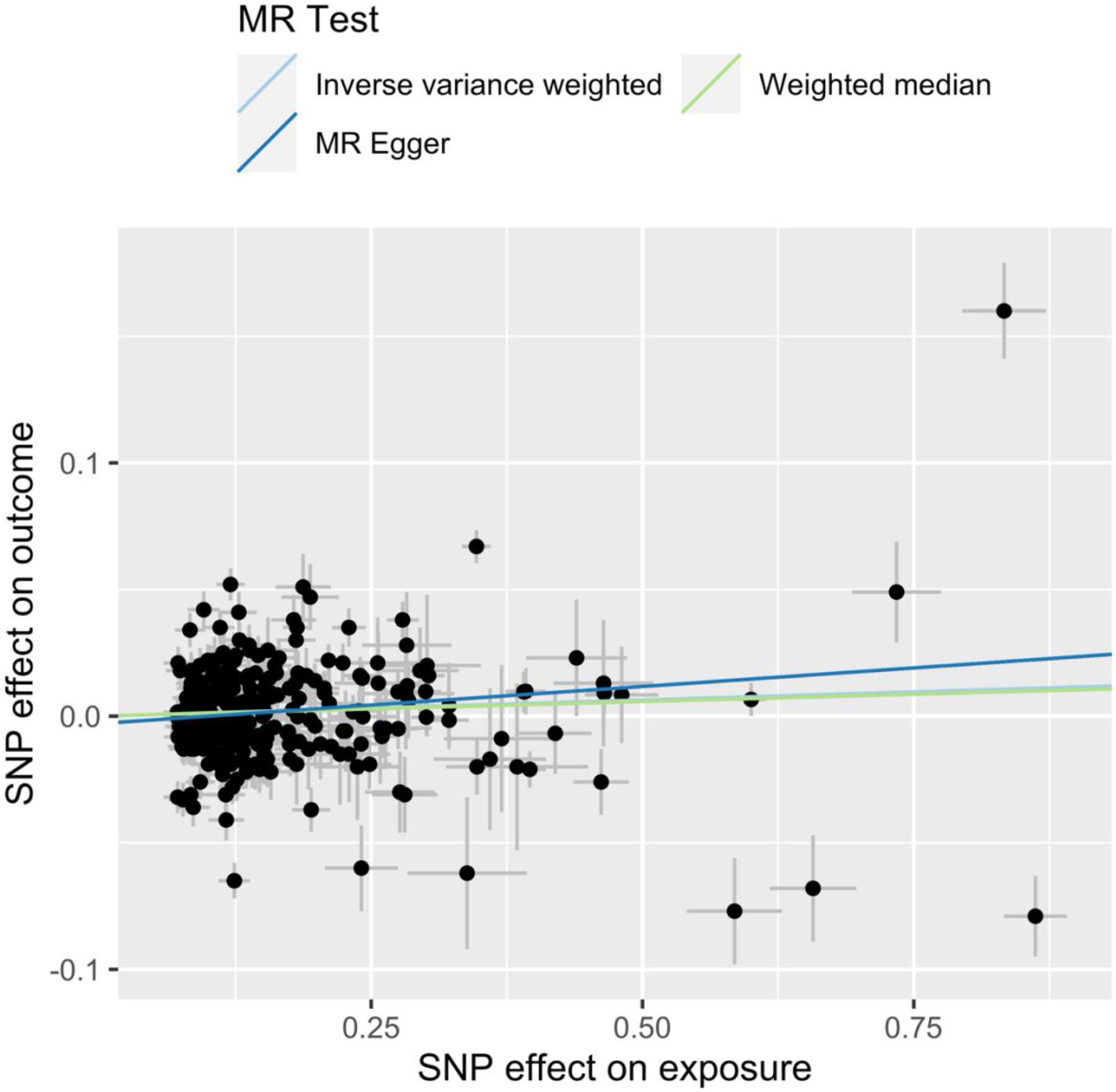
Scatter plots of the causal effect of continuous IGF-1 on type 2 diabetes for different MR tests. The x-axis is genetic association between SNPs and IGF-1 and the y-axis is genetic association between SNPs and type 2 diabetes. Analyses were conducted using the inverse variance weighed, weighted median and MR Egger methods. The slope of each line presents the estimated MR effect for each method.

**Table 3.** MR estimates from different methods of assessing the causal effect of 6 clusters on type 2 diabetes

| Clusters | Number of SNPs | Inverse variance weighted | Weighted median | MR Egger |
| --- | --- | --- | --- | --- |
|  |  | OR(CI) | OR(CI) | OR(CI) |
| Cluster 1 | 3 | 1.54 (1.43-1.65) | 1.54 (1.38-1.72) | 1.60 (1.00-2.57) |
| Cluster 2 | 41 | 1.03 (1.02-1.04) | 1.02 (1.01-1.04) | 1.01 (0.99-1.04) |
| Cluster 3 | 17 | 0.92 (0.91-0.94) | 0.93 (0.91-0.95) | 0.93 (0.90-0.96) |
| Cluster 4 | 4 | 0.62 (0.58-0.67) | 0.64 (0.56-0.73) | 0.52 (0.38-0.72) |
| Cluster 5 | 18 | 1.20 (1.18-1.22) | 1.21 (1.18-1.24) | 1.19 (1.14-1.23) |
| Cluster 6 | 3 | 0.80 (0.76-0.85) | 0.80 (0.75-0.86) | 0.91 (0.70-1.17) |

\*This table presents the odds ratio (OR) and 95% confidence interval (CI) of causal associations between 6 clusters and type 2 diabetes

**Figure 3.**
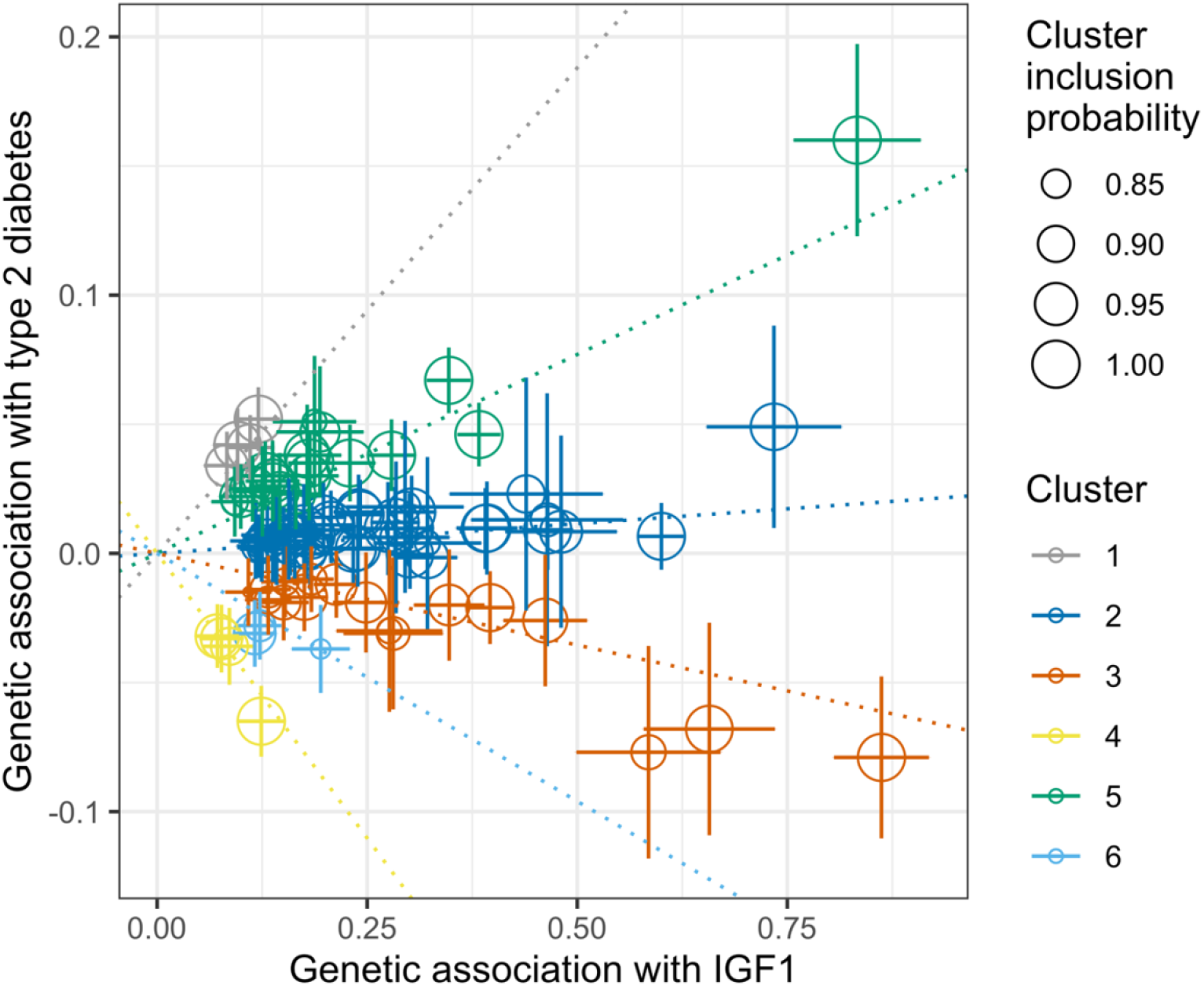
Scatter plots of the causal effect of continuous IGF-1 on type 2 diabetes for clustered MR analyses. The x-axis is genetic association between SNPs and IGF-1 and the y-axis is genetic association between SNPs and type 2 diabetes.

### Pathway analyses

After mapping the SNPs to genes, an overrepresentation analysis was performed using the KEGG and Reactome databases. We found that cluster 2, cluster 3 and cluster 4 mapped to specific pathways. Cluster 2 mapped to pathways related to systemic lupus erythematosus, metabolism of xenobiotics by cytochrome P450 and DNA/molecular integrity pathways (**Supplementary Figure 4**). Cluster 3 mapped to the pathways related to Janus kinase/signal transducers and activators of transcription (JAK/SAT), prolactin receptor signaling and GH receptor signaling (**Supplementary Figure 5**). Cluster 4 mapped to the pathways related to glycine, serine and threonine metabolism, choline catabolism and sulfur amino acid metabolism (**Supplementary Figure 6**). Clusters 1, 5 and 6 did not map to any specific pathway.

## Discussion and conclusion

In this study, we investigated the association of IGF-1 and type 2 diabetes using prospective multivariable-adjusted survival analyses followed by MR and clustered MR analyses in the UK Biobank. The results from the MR analyses showed that a genetically-influenced higher level of IGF-1 was associated with a higher risk of type 2 diabetes, which was in contrast to the result from the prospective analyses showing that a higher concentration of IGF-1 was associated with a lower risk of type 2 diabetes. Since the underlying individual genetic instruments showed a heterogeneous distribution of causal effect estimates, clustered MR identified 6 clusters of genetic instruments for IGF-1 with different associations with type 2 diabetes, which mapped to distinct molecular pathways. Collectively, our results indicate that the association between IGF-1 and the risk of developing type 2 diabetes is context-dependent.

Findings from other prospective studies regarding the association between IGF-1 and type 2 diabetes have been inconsistent. Our results from the prospective analyses were in line with another cohort study showing high levels of IGF-1 were associated with a lower risk of type 2 diabetes mellitus risk during 4.5 years of follow-up [5]. However, some nested case-cohort studies suggested there was no association between total IGF-1 levels and the risk of type 2 diabetes [28, 29]. In addition, a cohort study found that the association between free IGF-1 and type 2 diabetes was dependent on the level of insulin in women [30]. The inconsistency of these findings reinforces the notion that the association between IGF-1 and type 2 diabetes is context-dependent which is in line with the distinct biological mechanisms identified by the clustered MR analyses in our study.

Many studies found a J- or U-shaped association between IGF-1 and type 2 diabetes [5, 31] or insulin resistance [32]. For example, one study showed that individuals with both low- or high-IGF-1 levels were at increased risk of developing diabetes in a prospective cohort study [31]. Similarly, a U-shaped association between IGF-1 and measures of insulin resistance was found in a cross-sectional study in Danish adults [32]. Partly in line with these results, we observed a J-shaped relationship between IGF-1 and type 2 diabetes with particularly low levels of IGF-1 to be associated with an increased risk of developing type 2 diabetes. However, in the MR analyses of highest/lowest 20% IGF-1 and type 2 diabetes, we did not find indications that the association between genetically-influenced IGF-1 levels and type 2 diabetes was non-linear, as can be specifically observed in MR analyses in continuous IGF-1 where genetically-influenced low IGF-1 was associated with a lower risk of T2D and where genetically-influenced high IGF-1 was associated with a higher risk of T2D; effect sizes were similar from these analyses, but in opposite direction.

The result of MR analyses of continuous IGF-1 and type 2 diabetes in our study was supported by a recent publication showing higher levels of IGF-1 were causally associated with increased risk of developing type 2 diabetes [7]. However, we observed large between-SNP heterogeneity, and the causal estimates were not proportional to each other. We identified 6 main clusters of IGF-1 genetic instruments with distinct effects on type 2 diabetes by using clustered MR. After mapping the genetic instruments to genes and overrepresentation analysis using the KEGG and Reactome databases, several clusters were mapped to specific pathways.

Cluster 2 (in which higher genetically-influenced IGF-1 was associated with a higher risk of type 2 diabetes) mapped to multiple pathways all related to DNA/molecular integrity pathways. The association between the GH/IGF-1 axis and DNA damage and aging has long been observed [33]. For example, low IGF-1 expression was observed in the *Ercc1*^*-/-*^ mouse model, which also exhibits accelerated aging due to a genetic defect in DNA repair [34]. It has also been reported that developmental GH/IGF-1 deficiency is associated with increased cellular DNA repair capacity and upregulation of DNA repair-related genes [35]. These latter observations are consistent with genetically influenced and thus life-long higher IGF-1 levels and decreased DNA repair capacity. The association between the DNA damage response and type 2 diabetes is well described, yet poorly understood [36].

Cluster 3 (in which higher levels of genetically-influenced IGF-1 was associated with a lower risk of type 2 diabetes) was mapped to pathways related to GH receptor signaling. Dysregulation of GH receptor signaling and the GH-IGF-1 axis can lead to multiple diseases such as type 2 diabetes [37, 38]. Mice with liver IGF-1 deficiency had a fourfold increase in GH levels. Upon treatment with a GH antagonist these mice had decreased blood glucose and insulin levels and increased peripheral insulin sensitivity compared with mice with liver IGF-1 deficiency. These data indicate that the GH/IGF-1 axis plays a balancing role in insulin sensitivity and thus type 2 diabetes [39]. In addition, an epidemiological cross-sectional study showed that IGF-1 was associated with type 2 diabetes risk, but this association varied depending on the insulin levels: in individuals with low levels of insulin IGF-1 decreased type 2 diabetes risk and in individuals with high levels of insulin it increased type 2 diabetes risk [30].

Cluster 4 (in which high genetically-influenced IGF-1 levels were associated with a lower risk of type 2 diabetes) was mapped to pathways related to the metabolism of amino acids. Branched-chain amino acids have been associated with a higher risk of type 2 diabetes in several prospective studies [40, 41]. In addition, IGF-1 was able to affect protein metabolism by activating the mechanistic target of rapamycin (mTOR) pathway through phosphoinositide 3-kinase (PI3K) [42]. It was also found that muscle protein turnover was regulated by IGF-1 signaling via suppression of FoxO-regulated, autophagy-mediated protein degradation [43].

The main strength of our study is the extremely large sample size allowing stratification of the genetic instruments with ample statistical power. One limitation of the present study is that the IGF-1 level used was total IGF-1 concentration and not free IGF-1 (e.g., relative to the concentration of IGF binding proteins). Furthermore, the study was performed in a study of European-ancestry participants. Translation of the results to participants of non-European ancestry should be done with caution.

In conclusion, we found that a higher level of IGF-1 was causally associated with both a higher and a lower risk of development of type 2 diabetes and these inverse associations seem to be regulated by distinct biological mechanisms. Therefore, the total concentration of IGF-1 does not provide insight into the risk of developing type 2 diabetes.

## Data Availability

Data that support the findings of this study have been deposited in the UK Biobank under project number 22474. Data is available on request after approval of a research proposal by UK Biobank resources.

## Abbreviations

BMI: body mass index
GWAS: genome-wide association analyses
GH: growth hormone
IGF-1: insulin-like growth factor-1
MR: Mendelian Randomization
SNPs: single nucleotide polymorphisms

## Author Contributions

WW, EBT. KWvD, AB, DvH, RN: Substantial contributions to conception, design, acquisition of data, analyses and data interpretation; Drafting article and critically comment on the initial versions of the manuscript; Final approval of the manuscript before submission.

